# Burden of snakebite envenoming in northern Benin: A Retrospective Cross-Sectional Study in the Tchaourou District

**DOI:** 10.64898/2026.06.22.26356298

**Authors:** Enagnon Junior Juvénal Prince Honvou, Akpéna Ruth Esperencia Dèha, Jeannot Frejus Zinsou, Énagnon Parsifal Marie Alexandre Logbo, Francis Houeha, Michael Greenberg, Ralf Clemens, Ayola Akim Adegnika, Sue Ann Costa Clemens

## Abstract

**Background:** SBE is a significant public health problem, yet its incidence and associated disabilities and fatalities have been vastly underestimated.

**Objective:** To estimate the burden of snakebite envenoming in rural Tchaourou, Northern Benin.

**Methods:** A retrospective descriptive and analytical cross-sectional survey was conducted from 2018 to 2023 in the Tchaourou district. Data were collected through household interviews among participants of the ENABLE Lassa research program. Statistical analyses included incidence-rate calculations and multivariable logistic regression.

**Results:** Among 261 household respondents, 74 reported snakebites, yielding an incidence of 399 per 100,000 person-years [95% CI 314–501]. These 74 victims had a median age of 25.5 years and a male/female distribution of 55%:45% (41:33). Of the 74 victims, 32 (43%) reported that snakebites occurred while farming; and snakebites were most frequent during the rainy season (June and July) when farming activity was at its most intense. The most frequently bitten of the body was the foot or leg (74%), and the most reported symptoms included local swelling (78%), pain (78%), bleeding (66%), and headache (64%). The complications were reported by 22/74 (30%) victims. The risk of a snakebite complication was 3.0 times higher for female (14/33) than male victims (8/41; *p*=.032) and appeared to be higher when the first point of care was a traditional healer (12/44) or treatment at home (10/24) rather than at a health centre (0/6). Ultimately, only 18/74 (24%) victims attended a health centre.

**Conclusion:** Snakebite envenoming poses a significant health problem in Benin. Comprehensive strategies involving training of healthcare providers, community engagement, and improved immediate access to health centres should reduce morbidity.

**Author Summary:** Snakebite envenoming (SBE) is the second most deadly neglected tropical disease (NTD) and is a priority NTD for the World Health Organisation. In low-income tropical countries, SBE is a significant public health problem, yet its incidence and associated disabilities and fatalities have been vastly underestimated. We undertook a retrospective descriptive and analytical cross-sectional survey of 74 snakebite victims collected from 2018 to 2023 in the Tchaourou district of Benin, Africa. Our study suggested that the increased risk of snakebite comes from intensive farming in these tropical regions, and that previous underestimates may be related to only a minority of victims attending health centres where statistics are typically gathered. Moreover, our study suggested that improved immediate access to health centres should reduce morbidity, most notable for women.

## Introduction

Snakebite envenoming (SBE) is the second most deadly neglected tropical disease (NTD) (1), and is a priority NTD for the World Health Organisation (WHO) (2). Globally each year, 4.5 to 5.4 million people are bitten by snakes, 1.8 to 2.7 million are envenomed, and 81,000 to 138,000 die, with a case-fatality rate of 4 to 7% (2). Similarly, around 400,000 victims are left with disabilities, amputations, skin or muscle scars, brain, eye or kidney damage, social stigmatization and post-traumatic stress disorders (3–5).

In low-income tropical countries, SBE is a significant public health problem, yet its incidence and associated disabilities and fatalities have been vastly underestimated(2,3,6–9). In West Africa, the incidence of snakebites has been estimated at 8.9 to 93 per 100,000 person years, with a mortality rate of 0.5 to 5.9 per 100,000 person-years(8,9). In Benin, snakebites are most frequent in the departments of Borgou and Atacora, with an incidence of 144 per100,000 people-years and a case-fatality rate of 5.9% (10).

SBE imposes a substantial but often underestimated economic burden worldwide, as highlighted by a recent systematic review (11). In Southeast Asia, across seven countries including Malaysia, Thailand, Indonesia, the Philippines, Viet Nam, Lao PDR, and Myanmar, the total annual economic cost of snakebites was estimated at approximately USD 2.5 billion in 2019, representing about 0.09% of the region’s gross domestic product. Notably, more than 95% of this burden was attributable to premature mortality, underscoring the profound human and societal losses associated with snakebite-related deaths (12). In Benin, the use of antivenom has been shown to be highly cost-effective, with the cost per disability-adjusted life year averted estimated at approximately USD 83 (13). Furthermore, a hospital-based study in Benin reported a median direct hospital cost of snakebite management of around €168 per case, with costs varying considerably by clinical severity, ranging from approximately €31 for dry bites to €179 for cases complicated by external bleeding (14). Together, these findings illustrate not only the clinical but also the significant economic impact of SBE on affected households and health systems, particularly in resource-limited settings.

The district of Tchaourou is one of eight in the Borgou department and the largest district in Benin (15,16). The main activities in this district are farming and livestock breeding (15,16). In Benin, it has been shown that snakebite victims are often farmers and poor, and that there is a shortage of antivenomous serum in the areas concerned, with insufficient treatment equipment in health centres (17,18). The true disease burden of snakebite has been less well studied in Benin, and little attention has been paid to it by health authorities, relegating snakebite to a group of neglected public health problems (18).

WHO launched in 2019 the goal of reducing snakebites deaths and disabilities by 50% by 2030 (5). To measure impact and progress it is important to have baseline data at snakebite disease burden not only at national but also district level. Therefore, this study was conducted to: (i) determine the incidence of snakebites in rural areas of Tchaourou district (October 2018 to October 2023), (ii) analyse the profile of snakebite victims in Tchaourou communities, (iii) identify the factors associated with snakebites in Tchaourou communities.

## Methods

### Study area

This retrospective cross-sectional study was carried out in the district of Tchaourou, Benin (Figure 1). Tchaourou is the largest district in Benin, covering 7256 km², or around 6.5% of the national territory. Tchaourou borders Parakou, Pèrèrè and N’Dali to the north. To the east, it borders the Federal Republic of Nigeria, and to the west, the communes of Bassila and Djougou. Most roads are unpaved.

**Figure 1:**
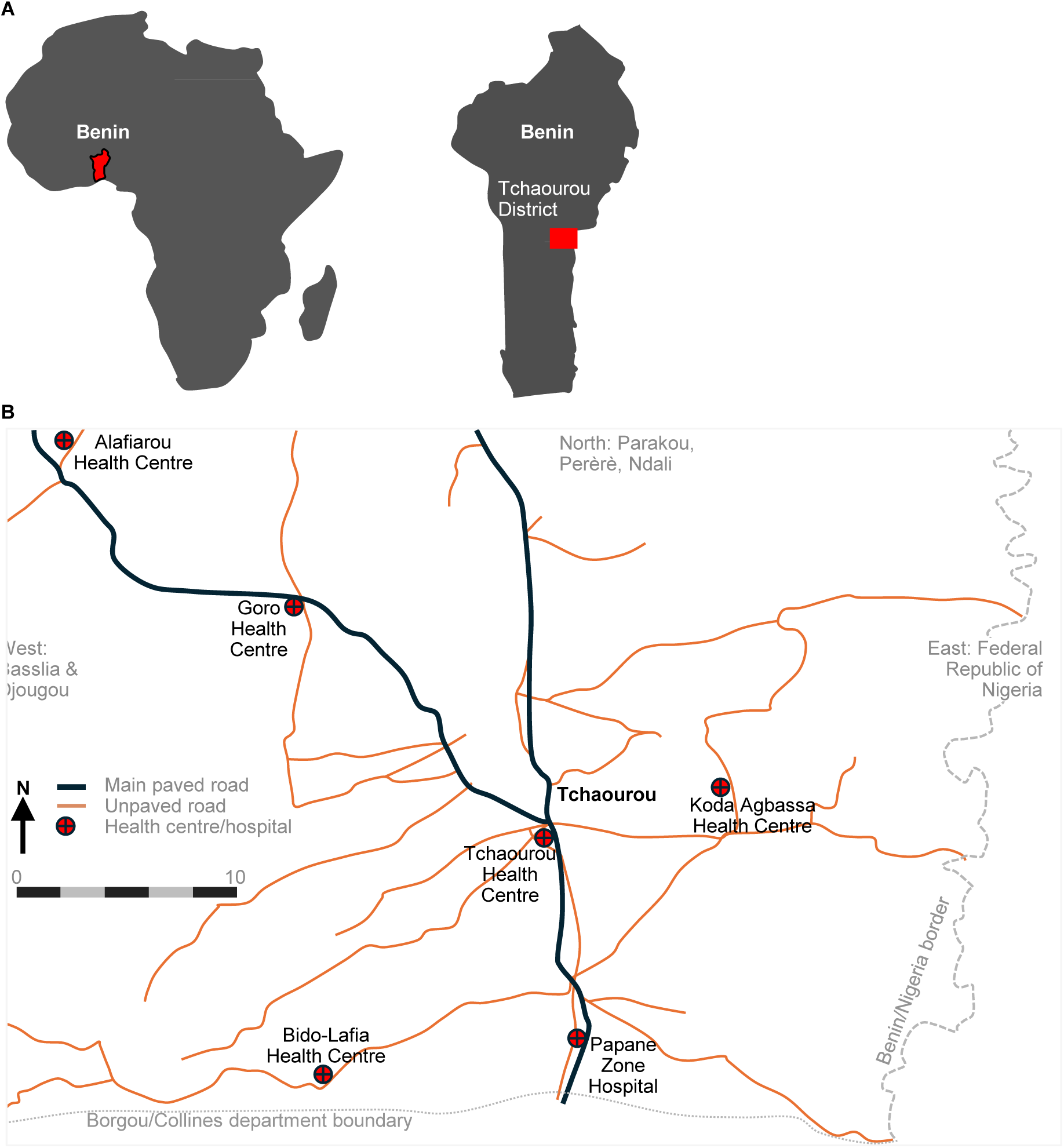
Geographical location of the district of Tchaourou in Benin. (A) The Tchaourou district in situated in the central region of Benin in Sub-Saharan Africa. (B) The district includes several health centres and is traversed by two principal paved roads.

In 2013, the population of Tchaourou was 223,118 according to the fourth General Census of Population and Housing (RGPH4) published in May 2013 (19), with an equal split between male and females. The agricultural population was 170,524. The main ethnic groups were the Baribas (40%), Peulhs (30%) and Nagots/Yorubas (6.0%). The population reported as being Muslim (66%), Catholic (15%), and, among others, animists (2.9%). The most common economic activities were classified under agriculture, livestock, fishing and hunting, which included 76% of the population (15,16). The climate is Sudanian, with a single rainy season. Annual rainfall varies between 900 and 1,300 mm. The rainy season begins in April and lasts around seven months. The average annual temperature is around 26°C, with a maximum of 32°C in March, dropping to around 23°C in December-January. Relative humidity varies between 30% and 70%. Four types of vegetation can be distinguished (15,16,19): the grassy savannah, tree and shrub savannah, wooded savannah.

### Data source

This study collected data from among households enrolled in the ENABLE 1.0 Lassa research program in Benin. This program was a prospective multi-site cohort study to estimate incidence of infection and disease due to Lassa fever virus in West African countries (20). The study localities were in well-defined geographical areas, including multiple rural and peri-urban localities, with healthcare facilities. To facilitate the acceptance of the study by the population, recruitment was done at household level rather than at individual level.

Two-stage cluster sampling method was used for the random selection of locations and households. A total of 34 villages in proportion to their population size, from a total of 36 villages in Tchaourou, Parakou districts was randomly selected. We used satellite imagery and a random selection algorithm to select 12 dwellings in each village plus six dwellings in reserve. The dwellings have been visited consecutively using a smartphone equipped with a GPS application until approximately 63 (+/- 3) participants were included.

The survey covered a five-year period from October 2018 to October 2023, and 264 households were interviewed.

The occurrence of snakebites and sociodemographics were considered for analysis. Data were collected during a face-to-face interview with the head of the household and/or its members, using a pre-established and validated questionnaire.

### Data analysis

All participating household members formed the analysis population. We validated the data by checking the completeness of the information collected using Kobocollect, a mobile data collection application, developed as part of the KoboToolbox platform and built on the Open Data Kit (ODK) framework. This was followed by data analysis using IBM SPSS Statistics 21 software.

We analysed the factors associated with the occurrence of complication by multivariable analysis using a top-down stepwise logistic model. For logistic regression, the dependent variable was crossed with the independent variables that were significant or had a *p* value ≤ 0.20 in univariate analysis. Statistical analysis used a *p* value of < 0.05 for all parameters to determine significant changes. Model fit was verified using the Lemeshow test. The odds ratios were presented with their 95% confidence intervals.

### Ethics and Data Privacy

The study was approved by the ethics committee of the University of Parakou (Comité Local d’Ethique pour la Recherche Biomédicale de l’Université de Parakou) N^0^: 0550/CLERB-UP/P/SP/R/SA. Rules of courtesy and privacy were respected towards each participant. Written informed consent was obtained from all adult participants. For participants younger than 18 years, written informed consent was obtained from a parent or legal guardian. In addition, written assent was obtained from children aged 10–17 years prior to participation. Written informed consent of the heads of household was obtained before the interview.

## Results

The study population of the 264 households included a total of 3706 participants with a median age of 18 years and a male/female distribution of 47%:53%. During the 5-year observation period from 2018 to 2023, 74 participants reported snakebites, representing an incidence rate of 399 bites per 100,000 person-year (95% CI 314–501). These 74 victims had a median age of 25.5 years and a male/female distribution of 55%:45% (41:33 victims; Table 1). The predominant age strata for male victims were 10 to 19 years (12), and 20 to 29 years (12), whereas for female victims it was ≥50 years (9). Most victims reported being bitten in the morning than other times of the day (28/74 [38%]), and during the rainy season between June and July (21/74 [28%]) than other months of the year (Table 2 and Figure 2). For male victims, the most prevalent activity in which the bite occurred was farming (21/41; 51%), whereas for female victims these activities were farming (11/33; 33%), walking outdoors (9/33;27%) and gathering wood (8/33; 24%) (Table 2).

**Figure 2:**
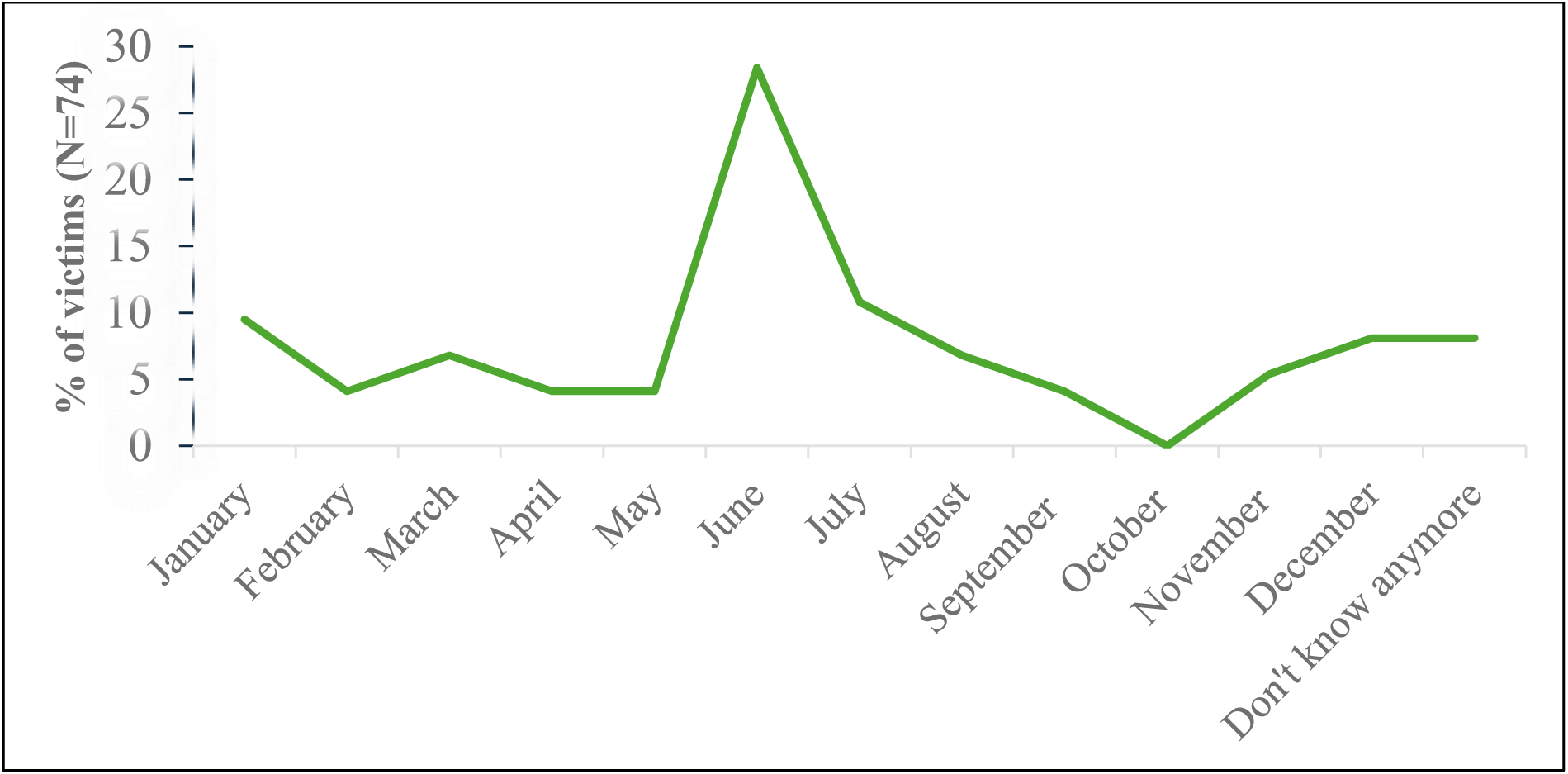
Distribution of snakebite victims by month of bite (N=74)

**Table 1:**
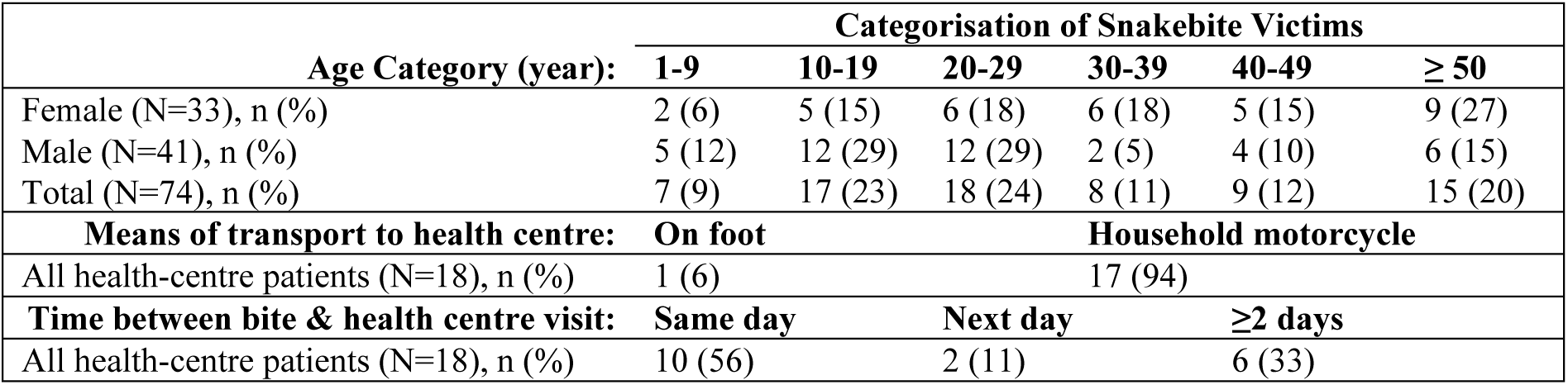
Categorisation of snakebite victims.

| **Age Category (year):** | **Categorisation of Snakebite Victims** |  |  |  |  |  |
| --- | --- | --- | --- | --- | --- | --- |
|  | **1-9** | **10-19** | **20-29** | **30-39** | **40-49** | **≥ 50** |
| Female (N=33), n (%) | 2 (6) | 5 (15) | 6 (18) | 6 (18) | 5 (15) | 9 (27) |
| Male (N=41), n (%) | 5 (12) | 12 (29) | 12 (29) | 2 (5) | 4 (10) | 6 (15) |
| Total (N=74), n (%) | 7 (9) | 17 (23) | 18 (24) | 8 (11) | 9 (12) | 15 (20) |
| **Means of transport to health centre:** | **On foot** |  |  | **Household motorcycle** |  |  |
| All health-centre patients (N=18), n (%) | 1 (6) |  |  | 17 (94) |  |  |
| **Time between bite & health centre visit:** | **Same day** |  | **Next day** |  | **≥2 days** |  |
| All health-centre patients (N=18), n (%) | 10 (56) |  | 2 (11) |  | 6 (33) |  |

**Table 2:** Distribution of victims by time of snakebite, activity at time of bite and first place of care.

|  | Number (Percentage) |  |  |
| --- | --- | --- | --- |
|  | Female (N=33) | Male (N=41) | Total (N=74) |
| **Time of snakebite** |  |  |  |
| Morning | 12 (36) | 16 (39) | 28 (38) |
| Noon | 2 (6) | 3 (7) | 5 (7) |
| Afternoon | 11 (33) | 13 (32) | 24 (32) |
| Evening | 8 (24) | 9 (22) | 17 (23) |
| **Activity at time of bite** |  |  |  |
| Farming | 11 (33) | 21 (51) | 32 (43) |
| Walking outdoors | 9 (27) | 7 (17) | 16 (22) |
| Gathering wood | 8 (24) | 0 (0) | 8 (11) |
| Livestock breeding | 1 (3) | 5 (12) | 6 (8) |
| Sleeping | 2 (6) | 2 (5) | 4 (5) |
| Hunting | 0 (0) | 3 (7) | 3 (4) |
| Playing outdoors | 1 (3) | 1 (2) | 2 (3) |
| While traveling | 0 (0) | 2 (5) | 2 (3) |
| Open defecation | 1 (3) | 0 (0) | 1 (1) |
| **First place of care** |  |  |  |
| Traditional healer | 22 (67) | 22 (54) | 44 (59) |
| Home | 8 (24) | 16 (39) | 24 (32) |
| Health centre | 3 (9) | 3 (7) | 6 (8) |

The snake responsible for the bite was identified by 53/74 (72%) of victims, with sand vipers (*Cerastes vipera)* being the most frequently reported type (16/53 [30%]; Table 3). The snakebites occurred on the foot or leg (58/74, 78%) and the finger, hand or forearm (16/74, 16%; Table 1). The most frequent symptoms reported by the snakebite victims were local swelling (58/74, 78%), local pain (58/74, 78%), local bleeding (49/74, 66%), and headaches (47/74, 64%; Figure 3).

**Figure 3:**
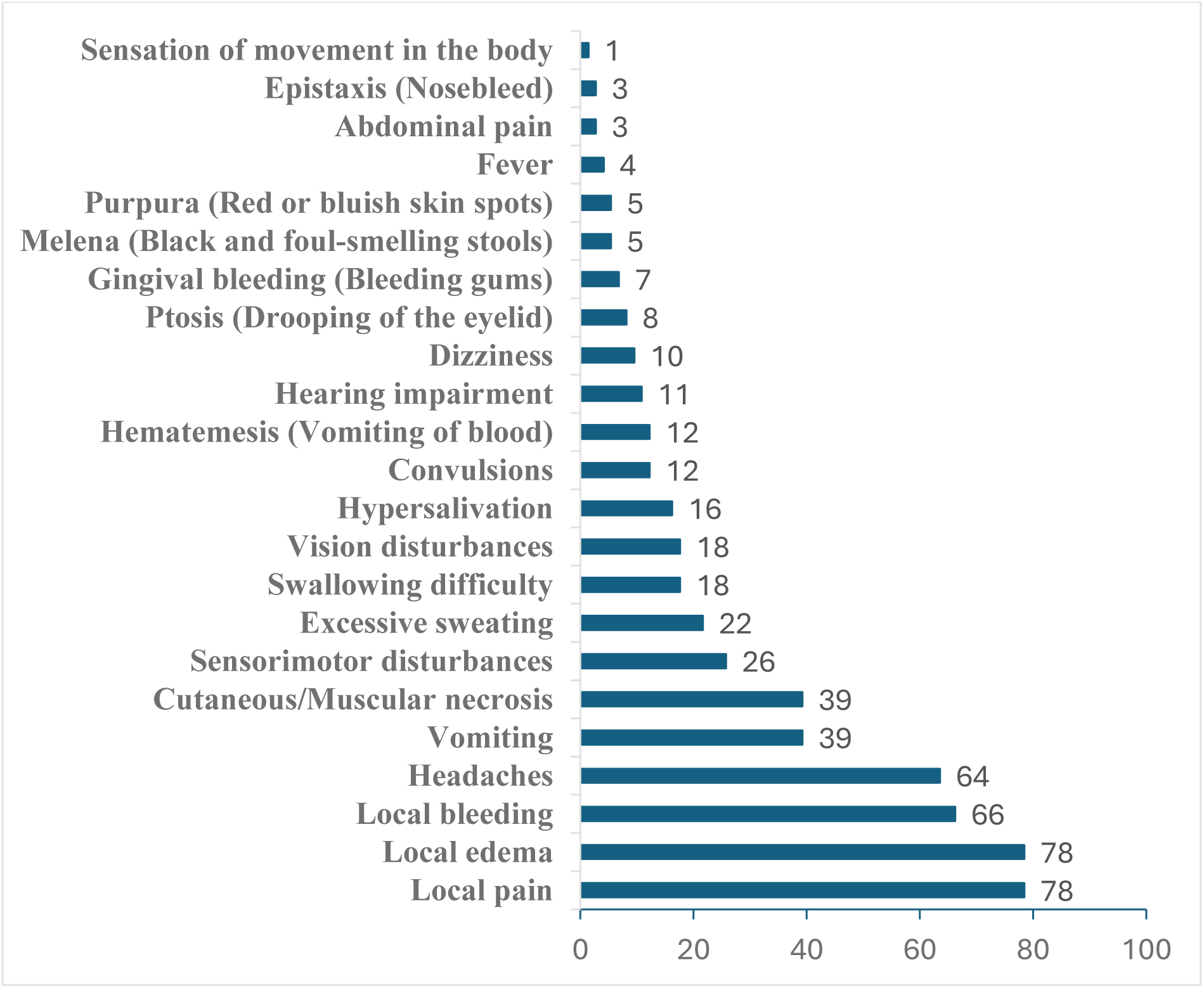
Distribution of symptoms reported by snakebite victims (N=74)

**Table 3:** Potential risk factors associated with snakebite complications.

|  | Complication<br>N | n (%) | Odds Ratio<br>(versus Reference [Ref]) | 95%<br>Confidence Interval | p-value |
| --- | --- | --- | --- | --- | --- |
| **Age (years)** |  |  |  |  |  |
| 1-9 | 7 | 1 (14) | Ref | - | - |
| 10-19 | 17 | 6 (35) | 3.27 | 0.31-33.94 | .320 |
| 20-29 | 18 | 4 (22) | 1.71 | 0.15-18.72 | .659 |
| 30-39 | 8 | 2 (25) | 2.00 | 0.14-28.41 | .609 |
| 40-49 | 9 | 4 (44) | 4.80 | 0.39-58.01 | .217 |
| ≥50 | 15 | 5 (33) | 3.00 | 0.27-32.20 | .364 |
| **Sex** |  |  |  |  |  |
| Male | 41 | 8 (20) | Ref | - | - |
| Female | 33 | 14 (42) | **3.03** | **1.07-8.56** | **.032** |
| **Snake type** |  |  |  |  |  |
| Echid | 7 | 0 (0) | Ref | - | .082 |
| Striking viper | 4 | 1 (25) | 0.77 | 0.07-7.91 | .831 |
| Elapsoide | 4 | 1 (25) | 0.77 | 0.07-7.91 | .831 |
| Naja | 3 | 1 (33) | 1.19 | 0.10-13.85 | .889 |
| Causas macule | 6 | 1 (17) | 0.47 | 0.05-4.41 | .465 |
| Black mamba | 5 | 4 (80) | **11.3** | **1.18-108.2** | **.011** |
| Horned viper | 6 | 2 (33) | 1.21 | 0.20-7.08 | .841 |
| Sand Viper | 16 | 5 (31) | 1.09 | 0.33-3.63 | .881 |
| **Body part bitten** |  |  |  |  |  |
| Hand/finger/forearm | 16 | 5 (31) | 1.09 | 0.33-3.63 | .881 |
| Foot/leg | 58 | 17 (29) | Ref | - | - |
| **1st care place** |  |  |  |  |  |
| Traditional healer | 44 | 12 (27) | Ref | - | - |
| Health centre | 6 | 0 (0) | - | - | .142 |
| Home | 24 | 10 (42) | 1.95 | 0.66-5.43 | .228 |
| **Surgery done** |  |  |  |  |  |
| No | 61 | 15 (25) | Ref | - | - |
| Yes | 13 | 7 (54) | **3.57** | **1.03-12.31** | **.036** |
| **Hospital stays** |  |  |  |  |  |
| 0-3 days | 10 | 3 (30) | Ref | - | - |
| 4-7 days | 7 | 2 (29) | 1.41 | 0.19-10.03 | .737 |
| More than 7 days | 1 | 1 (100) |  |  | .235 |
|  | Complication<br>N | n (%) | Odds Ratio<br>(versus Reference [Ref]) | 95%<br>Confidence Interval | p-value |
| **Age (years)** |  |  |  |  |  |
| 1-9 | 7 | 1 (14) | Ref | - | - |
| 10-19 | 17 | 6 (35) | 3.27 | 0.31-33.94 | .320 |
| 20-29 | 18 | 4 (22) | 1.71 | 0.15-18.72 | .659 |
| 30-39 | 8 | 2 (25) | 2.00 | 0.14-28.41 | .609 |
| 40-49 | 9 | 4 (44) | 4.80 | 0.39-58.01 | .217 |
| ≥50 | 15 | 5 (33) | 3.00 | 0.27-32.20 | .364 |
| **Sex** |  |  |  |  |  |
| Female | 33 | 14 (42) | **3.03** | **1.07-8.56** | **.032** |
| Male | 41 | 8 (20) | Ref | - | - |
| **Snake type** |  |  |  |  |  |
| Striking viper | 4 | 1 (25) | 0.77 | 0.07-7.91 | .831 |
| Echid | 7 | 0 (0) | Ref | - | .082 |
| Elapsoide | 4 | 1 (25) | 0.77 | 0.07-7.91 | .831 |
| Naja | 3 | 1 (33) | 1.19 | 0.10-13.85 | .889 |
| Causas macule | 6 | 1 (17) | 0.47 | 0.05-4.41 | .465 |
| Black mamba | 5 | 4 (80) | **11.3** | **1.18-108.2** | **.011** |
| Horned viper | 6 | 2 (33) | 1.21 | 0.20-7.08 | .841 |
| Sand Viper | 16 | 5 (31) | 1.09 | 0.33-3.63 | .881 |
| **Body part bitten** |  |  |  |  |  |
| Hand/finger/forearm | 16 | 5 (31) | 1.09 | 0.33-3.63 | .881 |
| Foot/leg | 58 | 17 (29) | Ref | - | - |
| **1st care place** |  |  |  |  |  |
| Health centre | 6 | 0 (0) | - | - | .142 |
| Home | 24 | 10 (42) | 1.95 | 0.66-5.43 | .228 |
| Traditional healer | 44 | 12 (27) | Ref | - | - |
| **Surgery done** |  |  |  |  |  |
| No | 61 | 15 (25) | Ref | - | - |
| Yes | 13 | 7 (54) | **3.57** | **1.03-12.31** | **.036** |
| **Hospital stays** |  |  |  |  |  |
| 0-3 days | 10 | 3 (30) | Ref | - | - |
| 4-7 days | 7 | 2 (29) | 1.41 | 0.19-10.03 | .737 |
| More than 7 days | 1 | 1 (100) |  |  | .235 |

The first point of care was similar for female and male victims, but overall, only 6/74 (8.1%) of snakebite victims were treated initially at a health centre, whereas 44/74 (59%) were treated initially by a traditional healer, and 24/74 (32%) were treated at home (Table 2). However, 18/74 (24%) of victims were treated ultimately at a health centre (Table 1), and 13/74 (18%) of victims had surgical interventions. Of those 18 victims, 10 attended the health centre on the same day, but eight victims arrived at least 2 days after the snake bite (Table 1). Seventeen were transported by a household motorcycle, and one by foot. Only one victim was hospitalised for more than 7 days (Table 3).

About a third of the snakebite victims (22/74, 30%) had complications (Table 3). These complications mainly concerned long-term pain (9/22, 41%), pain at the bite site during the rainy season (5/22, 23%), and physical problems affecting the ability to perform activities (5/22, 23%) (Figure 4). The risk of a complication from a snake bite was 3.0 times higher (p=0.032) for a female than a male victim (14/33 versus 8/41 victims; Table 3). The risk of a complication was 11 times higher (p=.011) with a black mamba bite than an echid bite (and concerned 3/4 versus 0/6 victims). The risk of complications was not associated with age or the site of the bite (notably, there were no bites to the face, which are considered to have higher complication rates). Although not statistically significant, the risk of complications may have been lower when the first point of care was at a health centre. No complication was reported by the six victims who were first given care at a health centre. By contrast, complications were reported by 12/44 (27%) victims who were first treated by a traditional healer and by 10/24 (42%) of victims who were treated at home. The risk of a complication was 3.6 times higher (p=.036) for victims who had surgery (7/13[54%]) than for those who did not (15/61[25%]). However, it was not documented whether the complications were present prior to or as a consequence of surgery.

**Figure 4:**
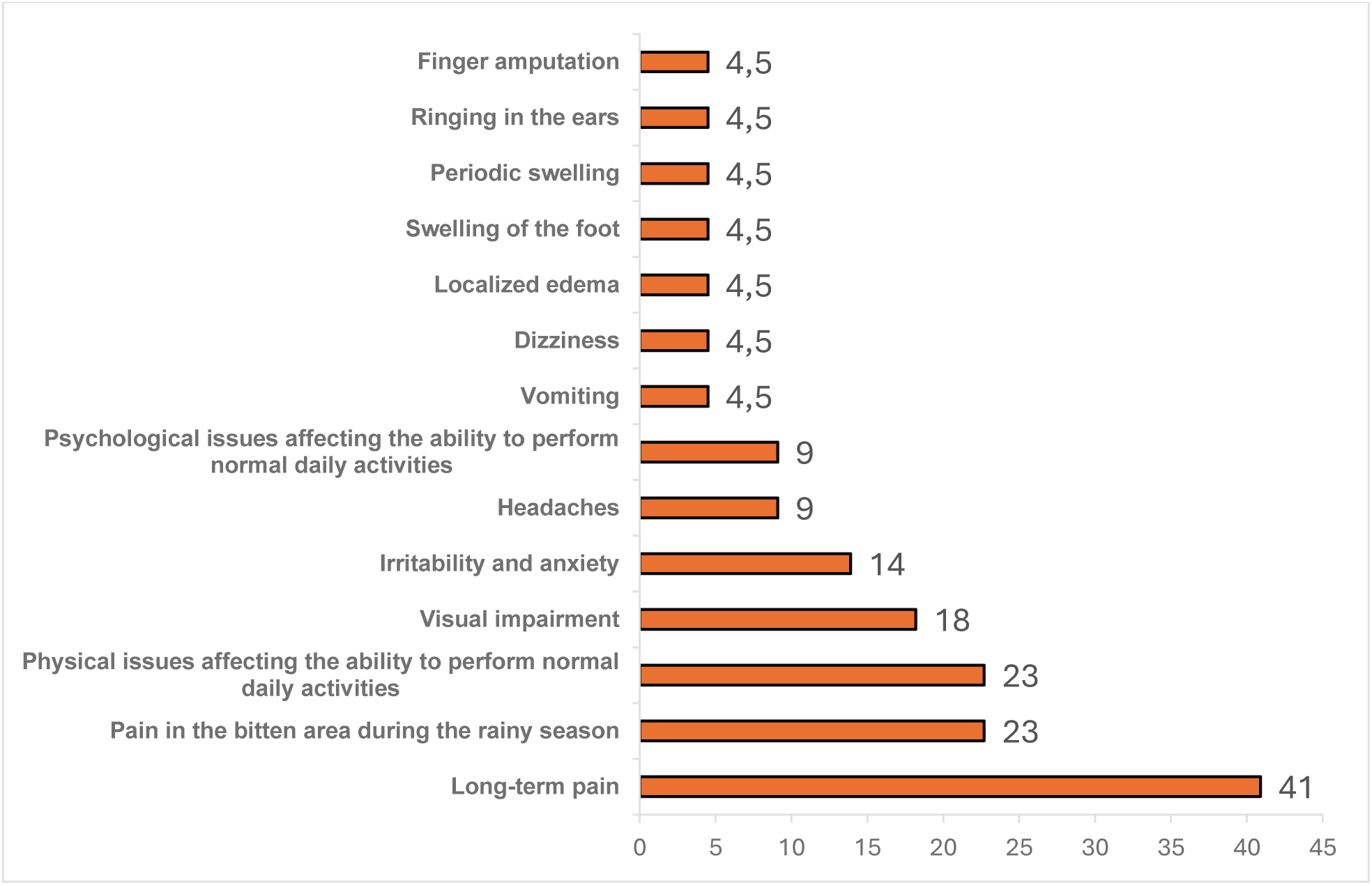
Complications in snakebite victims in the district of Tchaourou in 2023 by complications experienced after the bite (N=22)

## Discussion

Our study identified 74 snakebite victims in a major rural province in Benin between 2018 and 2023 representing an annual incidence rate of 399 per 100,000 person-years. Our study had several limitations: firstly, the study was retrospective and embedded in the Lassa epidemiology study, and therefore we could not assess in depth, the acute complications of a snakebite. Second, complications may have been missed in victims who were home treated and not followed up by a health centre. Third, no information was obtained on (i) whether a victim needed and received antivenom treatment, (ii) the victim’s family income, or (iii) the cost of the medical intervention. Nevertheless, our study points towards measures that could reduce the health burden caused by snakebite envenoming in Benin.

The incidence of snakebites of 399 per 100,000 person-years reported in this study was higher than earlier estimates in household surveys potentially because of the high degree of intensive farming in the Tchaourou province and therefore risk of exposure to snakes (19, 20), and stringent surveillance in the Lassa protocol. In other household surveys, the incidence was estimated as 144 per 100,000 person-years throughout Benin (10), 40 per 100,000 person-years in the plateau department of Benin (21), and 130 per 100,000 person-years in Burkina-Faso (22). Our estimate of snakebite incidence was also lower than estimates from hospital surveys in Benin (46 and 66 per 100,000 person-years (21)), which can be explained by the observation from our survey that only 24% of snakebite victims attended a health centre.

As in other studies (Benin and Ghana), adults appeared to be more at risk of snakebites than young children (6, 23). In our study, this may have been due to more adults being involved in farming (43% of bites occurred while farming), and by young adult males because 24 victims were male aged between 10 and 29 years). Moreover, as with other studies, the peak incidence of snakebites occurred during the rainy season, which coincides the peak farming period, for either planting, weeding, or harvesting (10,24,25). By contrast, female victims tended to be older than male victims, which may have been related to them also being involved in other activities in addition to farming, concurring with two other studies in Benin (10,18).

The types and frequencies of snakes responsible for the bites were typical of the region (26–28). The most frequent symptoms reported by the victims in the Tchaourou district were also typical for viper bites (local swelling [78%], pain [78%], and bleeding [66%], and headaches [64%]). However, the greatest risk of complications appeared to be with a black mamba bite.

The risk of a complication from a snake bite was 3.0 times higher for a female than a male victim, and this may have been related to the type of care received. The risk of complications appeared to be higher when the first point of care was a traditional healer or treatment at home rather than at a health centre. A factor behind these data may lie with insufficient means to attend health centres (lack of transport, time or finance), or lack of education or knowledge in best practice. Traditional treatments may have included, tourniquets, constricting bands and incisions for application of native herbs, which have no proven health benefit (10,21,23,29,30). By contrast, health benefits may have come from first aid measures such as victim reassurance, immobilization of the bitten limb with a splint or sling, and prompt visit to a health clinic, for potential treatment with an antivenom. The implementation of such approach has been achieved in Brazil, another country with a high SBE burden, where the majority of victims received medical assistance within 1 to 3 hours after being bitten (31). In one national study, medical assistance initiated within 6 hours was associated with about 42% lower odds of severe envenoming compared with later medical assistance (32).

## Data Availability

All relevant data underlying the findings of this study will be made publicly available in a recognized data repository prior to publication. The repository name and DOI will be provided upon acceptance of the manuscript

## Acknowledgements

We thank the CEPI and ENABLE 1.0 program team, the community health workers, and the residents of Tchaourou, Benin for their participation and support. Special thanks to Dr. Michael Greenberg and Celine Pompeia for their guidance throughout this research. Matthew Morgan (MG Science Communications, Belgium) provided scientific writing services.

## References

1. World Health Organization. Neglected tropical diseases [Internet]. Geneva: WHO; [cited 2022 Jan 11]. Available from: https://www.who.int/data/gho/data/themes/neglected-tropical-diseases

2. Gutiérrez JM, Calvete JJ, Habib AG, Harrison RA, Williams DJ, Warrell DA. Snakebite envenoming. Nat Rev Dis Primers. 2017;3:17063. doi:10.1038/nrdp.2017.63

3. Williams DJ, Faiz MA, Abela-Ridder B, Ainsworth S, Bulfone TC, Nickerson AD, et al. Strategy for a globally coordinated response to a priority neglected tropical disease: Snakebite envenoming. PLoS Negl Trop Dis. 2019;13(2):e0007059. doi:10.1371/journal.pntd.0007059

4. Kasturiratne A, Wickremasinghe AR, de Silva N, Gunawardena NK, Pathmeswaran A, Premaratna R, et al. The global burden of snakebite: A literature analysis and modelling based on regional estimates of envenoming and deaths. PLoS Med. 2008;5(11):e218. doi:10.1371/journal.pmed.0050218

5. World Health Organization. Snakebite: WHO targets 50% reduction in deaths and disabilities [Internet]. Geneva: WHO; 2019 [cited 2025 Sep 21]. Available from: https://www.who.int/news/item/06-05-2019-snakebite-who-targets-50-reduction-in-deaths-and-disabilities

6. Alcoba G, Ochoa C, Babo Martins S, Ruiz de Castañeda R, Bolon I, Wanda F, et al. Novel transdisciplinary methodology for cross-sectional analysis of snakebite epidemiology at national scale. PLoS Negl Trop Dis. 2021;15(2):e0009023. doi:10.1371/journal.pntd.0009023

7. Halilu S, Iliyasu G, Hamza M, Chippaux JP, Kuznik A, Habib AG. Snakebite burden in Sub-Saharan Africa: Estimates from 41 countries. Toxicon. 2019;159:1–4. doi:10.1016/j.toxicon.2018.12.002

8. Alcoba G, Chabloz M, Eyong J, Wanda F, Ochoa C, Comte E, et al. Snakebite epidemiology and health-seeking behavior in Akonolinga health district, Cameroon: Cross-sectional study. PLoS Negl Trop Dis. 2020;14(6):e0008334. doi:10.1371/journal.pntd.0008334

9. Habib AG, Kuznik A, Hamza M, Abdullahi MI, Chedi BA, Chippaux JP, et al. Snakebite is underappreciated: Appraisal of burden from West Africa. PLoS Negl Trop Dis. 2015;9(9):e0004088. doi:10.1371/journal.pntd.0004088

10. Chippaux JP. Snake bite epidemiology in Benin. Bull Soc Pathol Exot. 2002;95(3):172–174.

11. Patikorn C, Leelavanich D, Ismail AK, Othman I, Taychakhoonavudh S, Chaiyakunapruk N. Global systematic review of cost of illness and economic evaluation studies associated with snakebite. J Glob Health. 2020;10(2):020415. doi:10.7189/jogh.10.020415

12. Patikorn C, Blessmann J, Nwe MT, Tiglao PJG, Vasaruchapong T, Maharani T, et al. Estimating economic and disease burden of snakebite in ASEAN countries using a decision analytic model. PLoS Negl Trop Dis. 2022;16(9):e0010775. doi:10.1371/journal.pntd.0010775

13. Hamza M, Idris MA, Maiyaki MB, Lamorde M, Chippaux JP, Warrell DA, et al. Cost-effectiveness of antivenoms for snakebite envenoming in 16 countries in West Africa. PLoS Negl Trop Dis. 2016;10(3):e0004568. doi:10.1371/journal.pntd.0004568

14. Tourita N, Sodjinou N, Ouorou SA, Ganhouingnon É, Massougbodji A, Chippaux JP, et al. Evaluation of snakebite management cost at Saint Jean de Dieu Hospital in Tanguiéta, Benin. Med Trop Sante Int. 2024;4(4):mtsi.v4i4.2024.522. doi:10.48327/mtsi.v4i4.2024.522

15. Institut National de la Statistique et de la Démographie (INStaD). Cahier des villages et quartiers de ville: Département du Borgou [Internet]. Cotonou: INStaD; 2023 [cited 2026 Apr 2]. Available from: https://rgph5.instad.bj/wp-content/uploads/2023/03/BORGOU.pdf

16. Kora O. Monographie de la Commune de Tchaourou. Cotonou: Mission de décentralisation, Afrique Conseil; 2006.

17. Chippaux JP. Evaluation of the epidemiological situation and management of snakebite envenomation in francophone Sub-Saharan Africa. Bull Soc Pathol Exot. 2005;98:263–268.

18. Massougbodji A, Chobli M, Assouto P, Lokossou T, Sanoussi H, Sossou A, et al. Geoclimatology and severity of snake bite envenomations in Benin. Bull Soc Pathol Exot. 2002;95(3):175–177.

19. Institut National de la Statistique et de la Démographie (INStaD). RGPH4 – 2013 [Internet]. Cotonou: INStaD; 2013 [cited 2026 Mar 15]. Available from: https://rgph5.instad.bj/rgph4-2013/

20. Penfold S, Adegnika AA, Asogun D, Ayodeji O, Azuogu BN, Fischer WA, et al. A prospective, multi-site cohort study to estimate incidence of infection and disease due to Lassa fever virus in West African countries (Enable Lassa research programme): Study protocol. PLoS One. 2023;18(3):e0283643. doi:10.1371/journal.pone.0283643

21. Chippaux JP. Estimate of the burden of snakebites in Sub-Saharan Africa: A meta-analytic approach. Toxicon. 2011;57(4):586–599. doi:10.1016/j.toxicon.2010.12.022

22. Gampini S, Nassouri S, Chippaux JP, Semde R. Retrospective study on the incidence of envenomation and accessibility to antivenom in Burkina Faso. J Venom Anim Toxins Trop Dis. 2016;22:10. doi:10.1186/s40409-016-0066-7

23. Ceesay B, Taal A, Kalisa M, Odikro MA, Agbope D, Kenu E. Analysis of snakebite data in Volta and Oti Regions, Ghana, 2019. Pan Afr Med J. 2021;40:131. doi:10.11604/pamj.2021.40.131.28217

24. SK K, NR AV, Acharya A. Clinico-epidemiology and therapeutic outcome of snakebite in Konaseema region of Andhra Pradesh. J Evid Based Med Healthc. 2016;3(29):1314–1316. doi:10.18410/jebmh/2016/302

25. Ooms GI, van Oirschot J, Waldmann B, Okemo D, Mantel-Teeuwisse AK, van den Ham HA, et al. The burden of snakebite in rural communities in Kenya: A household survey. Am J Trop Med Hyg. 2021;105(3):828–836. doi:10.4269/ajtmh.21-0266

26. World Health Organization. Guidelines for the prevention and clinical management of snakebite in Africa [Internet]. Geneva: WHO; [cited 2022 Jan 11]. Available from: https://www.who.int/publications/i/item/9789290231684

27. Visser LE, Kyei-Faried S, Belcher DW. Protocol and monitoring to improve snakebite outcomes in rural Ghana. Trans R Soc Trop Med Hyg. 2004;98(5):278–283. doi:10.1016/S0035-9203(03)00065-8

28. Trape JF, Pison G, Guyavarch E, Mane Y. Mortality from snake bites and other animal-related injuries in eastern Senegal. Bull Soc Pathol Exot. 2002;95(3):154–156.

29. Meshram RM, Bokade CM, Merchant S, Bhongade S. Clinical profile and outcome of snake bite in children. Int J Contemp Pediatr. 2017;4(3):910–914. doi:10.18203/2349-3291.ijcp20171697

30. Michael GC, Bala AA, Mohammed M. Snakebite knowledge assessment and training of healthcare professionals in Asia, Africa, and the Middle East: A review. Toxicon X. 2022;16:100142. doi:10.1016/j.toxcx.2022.100142

31. Oliveira HF, Barros RM, Pasquino JA, Peixoto LR, Sousa JA, Leite RS. Snakebite cases in municipalities of Paraíba State, Brazil. Rev Soc Bras Med Trop. 2013;46(5):617–624. doi:10.1590/0037-8682-0130-2013

32. Mise YF, Lira-da-Silva RM, Carvalho FM. Time to treatment and severity of snake envenoming in Brazil. Rev Panam Salud Publica. 2018;42:e52. doi:10.26633/RPSP.2018.52

